# Patient-Specific Adaptations in ERAS for High-Altitude Laparoscopic Cholecystectomy: The PAERS Hypothesis

**DOI:** 10.64898/2026.08.21.26360905

**Authors:** Zhongfeng Dang, Jianduojie Dan, Wei Su, Guoliang Ren, Zhiqiang Wang, Yabing Ma, Shengmei Li, Dongde Ji, Liansheng Li, Junlin Gao

**Author notes:** Co-first authors (Zhongfeng Dang, Jianduojie Dan). Co-second authors (Wei Su, Guoliang Ren, Zhiqiang Wang, Yabing Ma). **Corresponding author**: Zhongfeng Dang, Department of Hepatobiliary Surgery, Qinghai Red Cross Hospital, Xining, Qinghai 810000, China. **Co-corresponding author**: Junlin Gao, Department of Hepatobiliary Surgery, Qinghai Red Cross Hospital, Xining, Qinghai 810000, China.

## Abstract

**Background:** ERAS protocols reduce hospital stay by 1.88 days and complications by 29% globally, but their “one-size-fits-all” paradigm—validated at sea level—may fail at high altitude where chronic hypoxia and population-specific genetic adaptations remodel baseline physiology. No study has quantified ERAS effect weight shifts at high altitude or proposed a theoretical model to explain the gap.

**Objectives:** To evaluate three dimensions of plateau ERAS remodeling—(i) risk factor weight shift, (ii) traditional marker failure, (iii) genetic background modification—and propose the PAERS (Plateau Adaptation–ERAS Remodeling Syndrome) risk stratification model tailored to altitude.

**Methods:** Retrospective cohort of 612 adults undergoing elective laparoscopic cholecystectomy (2018–2023) at Qinghai Red Cross Hospital (2260 m). Three analytical tiers: (1) multivariable regression comparing risk factor coefficients against plain-altitude benchmarks; (2) restricted cubic spline and interaction modeling for Hb, SpO₂, and LOS; (3) inferential genetic modifier analysis using population-level EPAS1 carrier rates. Primary outcomes: LOS and complication rate.

**Results:** Three-dimensional shift was observed: **(1) Weight Remodeling**: BMI replaced sex as primary risk factor (OR = 1.86, P < .001), surgeon variability amplified (F = 6.33 vs plain benchmark 2–4, an ∼58% increase in F-statistic ratio, P < .001); **(2) Marker Failure**: Hb showed J-type relationship with LOS (Hb × SpO₂ interaction β = −0.0095, P = .009), with effect reversal across SpO₂ strata (”Plateau Hemoglobin Paradox”); **(3) Genetic Modification (population-level inference)**: ∼70% EPAS1 carrier rate (range 57–85% across studies) suggests HIF-2α pathway is a baseline modifier that must be accounted for. Three falsifiable predictions were proposed.

**Conclusions:** High-altitude ERAS faces three challenges: effect weight remodeling, biomarker failure, and genetic background calibration. The PAERS hypothesis proposes an integrated risk stratification model—shifting from “one-size-fits-all” to altitude-aware, patient-specific protocols.

## 1. INTRODUCTION

The global implementation of Enhanced Recovery After Surgery (ERAS) protocols has demonstrated benefits in reducing hospital length of stay (LOS) and postoperative complications across diverse surgical specialties. A 2024 meta-analysis by Sauro and colleagues, synthesizing data from 74 randomized controlled trials enrolling 9076 participants, confirmed that ERAS reduces LOS by a weighted mean of 1.88 days (95% CI: 0.95–2.81) and overall complication risk by 29% (risk ratio 0.71; 95% CI: 0.59–0.87). [1] These benefits have led to widespread adoption endorsed by the ERAS Society and national guideline bodies worldwide.[2,3] The evidence base underpinning every component of current ERAS pathways—from risk stratification thresholds to intervention effect sizes—derives exclusively from studies conducted at or near sea level. This body of work implicitly assumes a “universal ERAS paradigm”: that the relative weights of perioperative risk factors, the prognostic value of clinical biomarkers, and the optimal intensity of interventions remain constant across all populations and environments. Whether this implicit assumption holds in high-altitude settings, where chronic hypoxia and population-specific genetic adaptations remodel baseline physiology, remains an unanswered question of direct clinical relevance to over 140 million permanent high-altitude residents globally.

High-altitude environments (altitude > 2500 m) differ physiologically from sea-level settings, beyond simple atmospheric variation. At 2260 m, atmospheric oxygen partial pressure is approximately 75% of sea-level values, driving chronic hypoxic adaptation across multiple organ systems.[4] Highland populations, particularly Tibetans, carry near-universal adaptive genetic variants in EPAS1 (encoding HIF-2α) and EGLN1, with carrier rates approaching 70%.[5–7] These variants regulate erythropoiesis, inflammation, angiogenesis, and metabolic reprogramming—all central to perioperative recovery. Several studies have confirmed that ERAS protocols are feasible at high altitude, demonstrating reduced LOS and complications compared to conventional care.[8–10] However, these studies adopted plain-validated ERAS pathways without questioning whether the effect weights of individual components remain valid in genetically adapted, chronically hypoxic populations. The clinical paradox is evident: ERAS is feasible at high altitude, but 24-hour discharge rates remain approximately 50% versus > 80% at sea level, and complication rates are 12–18% versus < 5% in plain settings.[8,9] This gap between feasibility and optimality has not been mechanistically explained.

The existing knowledge gap can be decomposed into three testable dimensions. **(i) The Effect Weight Shift Gap**: Do risk factor rankings derived from plain-altitude ERAS models—typically dominated by sex, age, and ASA grade—remain valid at high altitude? Preliminary data from our group suggest BMI may supersede sex as the primary risk factor, but the magnitude of this shift remains unquantified.[11] **(ii) The Marker Failure Gap**: Are traditional ERAS risk stratification biomarkers—hemoglobin (Hb), albumin, and ASA grade—still prognostically useful at high altitude? The non-linear relationship between Hb and oxygen-carrying capacity under chronic hypoxia raises the specific hypothesis that Hb may exhibit paradoxical associations with surgical outcomes rather than the monotonic protective relationship assumed in plain protocols.[12] **(iii) The Genetic Modifier Gap**: How does the near-universal presence of hypoxia-adaptive genetic variants (EPAS1, EGLN1) in highland populations modify surgical stress responses? Current ERAS risk models entirely omit genetic background, yet HIF-2α pathway activation is known to regulate erythropoiesis, inflammation, angiogenesis, and metabolic reprogramming.[5,13]

This study aims to evaluate these three dimensions and propose the **PAERS (Plateau Adaptation–ERAS Remodeling Syndrome) hypothesis**: in high-altitude populations with chronic hypoxic adaptation and elevated hypoxia-adaptive genetic variant carrier rates, the translation of plain-validated ERAS protocols is subject to three remodeling effects requiring adjustment: (1) **Risk Weight Remodeling**, where baseline risk factor ranking and effect sizes shift due to hypoxic amplification of specific physiological vulnerabilities; (2) **Biomarker Functional Remodeling**, where dose-response curves of traditional plain biomarkers change shape—including J-type nonlinearity and SpO₂-dependent effect reversal—rendering plain-derived thresholds invalid; and (3) **Genetic Background Modification**, where population-level HIF pathway activation constitutes a baseline modifier that must be accounted for. The observed gap between ERAS feasibility and optimality at high altitude requires an explanatory framework that accounts for the divergence from plain-validated effect sizes—a divergence that single-dimensional explanations cannot reconcile. We conducted a retrospective cohort study of 612 patients undergoing laparoscopic cholecystectomy (LC) at a high-altitude center, integrating findings from four interconnected analyses—surgical efficiency heterogeneity, epidemiological risk factor remodeling, health economics cost-control modeling, and the Hb–SpO₂ interaction (the “Plateau Hemoglobin Paradox”)—to provide the first multi-dimensional empirical test of the PAERS framework.[11,14–16]

## 2. METHODS

### 2.1 Study Design and Setting

This was a hypothesis-generating, single-center retrospective cohort study conducted at Qinghai Red Cross Hospital (Xining, China; altitude 2260 m). The study period spanned January 1, 2018, to December 31, 2023. The Institutional Review Board approved the study (No. LW-2026-73) with a waiver of informed consent. This study followed the STROBE reporting guidelines.[17] The PAERS hypothesis and its three testable dimensions were formulated post hoc through integration of four interconnected analyses conducted by our group (surgical efficiency, epidemiological, health-economic, and biomarker-outcome). All subgroup, sensitivity, and post-hoc power analyses were exploratory and interpreted with nominal P values, consistent with STROBE guidance for hypothesis-generating research.

### 2.2 Participants and Eligibility Criteria

#### Inclusion

(1) age ≥ 18 years; (2) elective laparoscopic cholecystectomy for symptomatic cholelithiasis or gallbladder polyps; (3) ASA physical status I–III; (4) complete preoperative Hb, SpO₂, and LOS data; (5) surgery performed under a standardized ERAS protocol.

#### Exclusion

(1) conversion to open cholecystectomy; (2) severe preoperative anemia (Hb < 60 g/L); (3) Hb > 250 g/L; (4) severe cardiopulmonary comorbidity (NYHA class III–IV heart failure, severe COPD, or pulmonary hypertension); (5) perioperative blood transfusion; (6) postoperative LOS > 30 days; (7) incomplete data.

Of 742 patients screened, 612 were included in the final analytic cohort (Figure 1). This cohort overlaps with four interconnected analyses from our group, each focusing on independent scientific questions: surgical efficiency heterogeneity (n = 591, top 7 surgeons only),[14] epidemiological risk factor remodeling (n = 605),[15] health economics cost-control modeling (n = 605),[16] and the Hb–SpO₂ interaction (n = 612).[11] Sample size differences arise from analysis-specific exclusion criteria. The present study uses the maximum available sample (N = 612) and integrates all four sets of findings.

**Figure 1.**
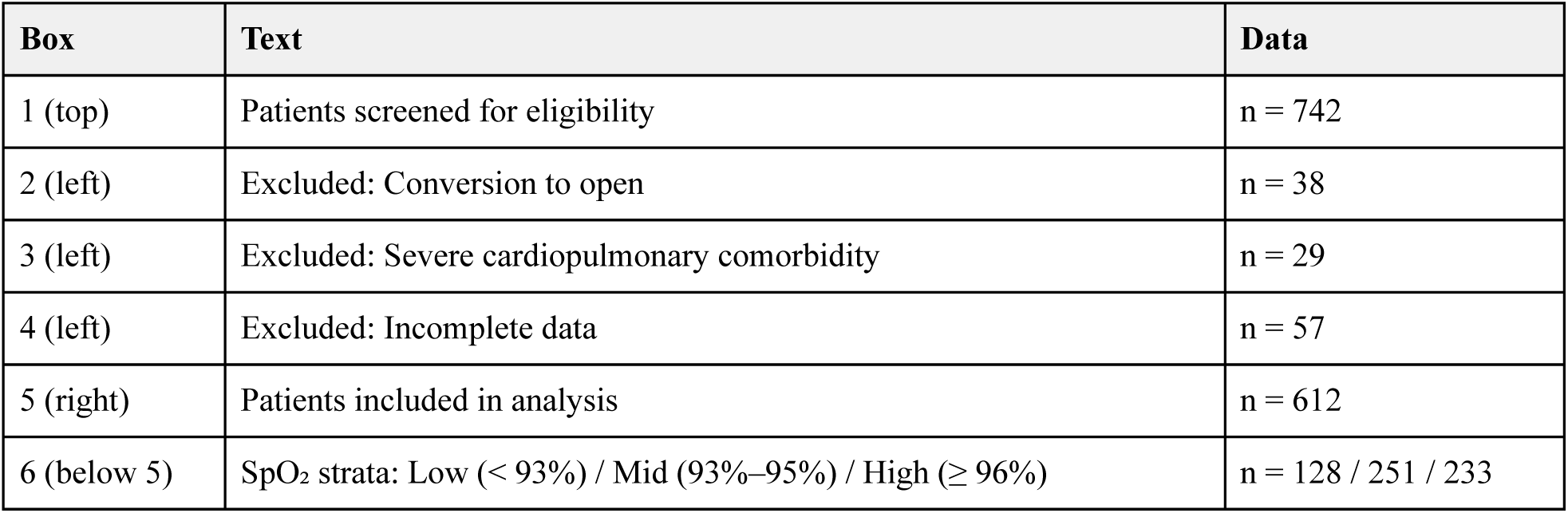
Study Flow Diagram.

### 2.3 Variables

#### Primary outcome

Postoperative length of stay (LOS), defined as days from surgery to discharge. Discharge was determined by standardized ERAS criteria.[2]

#### Secondary outcomes

Complication rate (Clavien-Dindo ≥ grade II), operative time (skin incision to closure, minutes), total hospitalization cost (CNY), and 30-day readmission rate.

**Exposure variables** were organized into three dimensions:

##### Dimension 1 — Baseline risk factors (for weight shift analysis)

Age (years), sex (female vs male), BMI (kg/m²; categorized as underweight < 18.5, normal 18.5–23.9, overweight 24.0–27.9, obese ≥ 28 per Chinese guidelines).

##### Dimension 2 — Clinical biomarkers (for marker failure analysis)

Preoperative Hb (g/L; measured within 72 hours before surgery; Sysmex XN-9000), categorized as < 180 vs ≥ 180 g/L for subgroup analyses. Preoperative SpO₂ (%; fingertip pulse oximetry, Masimo Radical-7; measured at rest, supine, on the day of surgery), stratified as low (< 93%), mid (93%–95%), and high (≥ 96%).[12] Serum albumin (g/L) and ASA grade (I/II/III).

##### Dimension 3 — Surgical and environmental variables

Surgeon identity (7 high-volume surgeons, de-identified as Surgeon A–G), operative time, and season (winter vs non-winter).

##### Genetic modifier variable (inferential)

Individual genotyping was not performed. Population-level EPAS1 adaptive variant carrier rates (∼70%) were inferred from published Tibetan plateau genomics literature and our group’s prior work in the same geographic population.[5–7,18] All genetic modification analyses are explicitly labeled as population-level inferences, not individual-level empirical findings.

### 2.4 Statistical Analysis

#### Descriptive statistics

Continuous variables as mean ± SD or median (IQR); categorical variables as frequency (%). Between-group comparisons used one-way ANOVA with Bonferroni correction, χ², or Fisher’s exact test.

#### Tier 1 — Effect Weight Shift

(a) Multivariable logistic regression for cholecystitis progression with BMI, sex, age, blood pressure, and diagnosis type as covariates; ORs compared against plain-altitude benchmarks (sex OR ≈ 1.5–2.0; BMI OR ≈ 1.2–1.4).[19,20] (b) One-way ANOVA for surgeon variability in operative time (F vs plain benchmark 2–4).[21,22] (c) Multivariable linear regression for ln(total cost), comparing LOS and operative time β coefficients against plain benchmarks.

#### Tier 2 — Marker Failure

(a) Restricted cubic spline (RCS) regression for the Hb–LOS relationship.[23] (b) Multivariable linear regression with Hb × SpO₂ interaction term (full equation in Supplementary Material S8). Effect reversal was defined post hoc as β₁ and (β₁ + β₃) having opposite signs with β₃ reaching P < .05. (c) Model fit comparison: “plain model” (BMI + Hb + albumin + ASA) vs “enhanced model” (plain model + SpO₂ + Hb × SpO₂ interaction), compared using R², LRT, ΔAIC, and Cohen’s f².

#### Tier 3 — Genetic Modification (Inferential, population-level)

A three-step inference chain was constructed: (1) EPAS1+ carrier rate (∼70%, range 57–85% across Tibetan plateau genomics studies[36–39]) → HIF-2α pathway baseline activation; (2) HIF-2α → Hb set-point down-regulation, inflammatory/vascular/metabolic baseline shift; (3) baseline shift → adjustment of surgical stress response. Three falsifiable predictions (F1–F3) were formulated for prospective Phase 3 validation.

#### HERS Score (Conceptual, see Supplementary Material S1)

A High-altitude ERAS Score (HERS) was constructed as a conceptual model. Given its conceptual-only status (AUC = 0.69, insufficient discrimination, not externally validated), it is presented for illustrative purposes only and must not be used for clinical decision-making. Detailed construction is provided in Supplementary Material S1.

#### Sensitivity analyses

Five computational robustness analyses from our companion study[11] were referenced (see Supplementary Material S8 for details).

#### Post-hoc power

Achieved power was calculated using the non-central F distribution (α = 0.05, df₁ = 1, df₂ = 604).

Analyses used Python 3.12 (pandas, numpy, scipy, statsmodels, scikit-learn). All P values were two-sided; 95% CIs are reported.

## 3. RESULTS

### 3.1 Study Population

Of 742 patients screened, 612 were included (Figure 1). Mean age was 43.5 (SD 11.9) years; 65.8% were female (Table 1). Mean preoperative Hb was 151.4 (SD 20.7) g/L, with 50 patients (8.2%) having Hb ≥ 180 g/L. Mean SpO₂ was 94.6% (SD 2.3%). SpO₂ strata comprised 128 (20.9%) low (< 93%), 251 (41.3%) mid (93%–95%), and 233 (37.9%) high (≥ 96%). Age differed significantly across strata (P < .001); sex and BMI were comparable (P = .66 and P = .11). Mean LOS was 1.6 (SD 0.7) days.

**Table 1.** Baseline Characteristics by SpO₂ Stratum (N = 612)

| Variable | Overall (N = 612) | Low SpO<sub>2</sub> < 93%<br>(n = 128) | Mid SpO<sub>2</sub> 93%–95% (n = 251) | High SpO<sub>2</sub> ≥ 96%<br>(n = 233) | P |
| --- | --- | --- | --- | --- | --- |
| Age, mean (SD), y | 43.5 (11.9) | 47.1 (12.3) | 43.5 (11.9) | 41.6 (11.2) | < .001 |
| Female sex, No. (%) | 403 (65.8) | 89 (69.5) | 164 (65.3) | 150 (64.4) | .66 |
| BMI, mean (SD), kg/m<sup>2</sup> | 24.2 (3.8) | 24.6 (3.8) | 24.4 (3.8) | 23.8 (3.7) | .11 |
| Hemoglobin, mean (SD), g/L | 151.4 (20.7) | 153.4 (19.6) | 152.1 (20.7) | 149.5 (21.1) | .18 |
| Hb ≥ 180 g/L, No. (%) | 50 (8.2) | 9 (7.0) | 25 (10.0) | 16 (6.9) | .40 |
| SpO<sub>2</sub>, mean (SD), % | 94.6 (2.3) | 91.2 (0.9) | 94.2 (0.8) | 96.8 (1.0) | < .001 |
| Operative time, mean (SD), min | 48.3 (18.8) | 50.0 (17.3) | 47.3 (14.9) | 48.5 (22.8) | .42 |
| Winter season, No. (%) | 167 (27.3) | 43 (33.6) | 68 (27.1) | 56 (24.0) | .17 |
| LOS, mean (SD), d | 1.6 (0.7) | 1.8 (1.0) | 1.6 (0.5) | 1.6 (0.7) | .043 |

### 3.2 Dimension 1: Risk Factor Weight Remodeling

#### BMI replaced sex as the primary risk factor

In multivariable logistic regression for cholecystitis progression (n = 605), obesity (BMI ≥ 28) was the sole independent risk factor (OR = 1.86; 95% CI: 1.32–2.62; P < .001), while sex showed no independent effect (OR = 1.08; 95% CI: 0.75–1.57; P = .668). The BMI × sex interaction was non-significant (P = .892), confirming that BMI’s effect does not vary by sex—it has simply superseded sex as the dominant risk factor.[15] Against published plain-altitude benchmarks (sex OR ≈ 1.5–2.0; BMI OR ≈ 1.2–1.4),[19,20] the high-altitude pattern shows a complete rank reversal: BMI effect increased by approximately 43%, while sex effect attenuated to statistical null.

In LOS regression (n = 612), BMI was the strongest baseline predictor (β = +0.028 d/kg/m²; P < .001), with female sex only marginally significant (β = +0.149 d; P = .044). The model (R² = 0.047; F = 5.04; P < .001) identified BMI as the dominant risk factor for prolonged hospital stay.[11]

#### Surgeon variability was amplified at high altitude

Among 7 surgeons (n = 591), operative time ANOVA yielded F = 6.33 (P < .001),[14] compared to a plain-altitude benchmark range of F ≈ 2–4.[21,22] This represents a 58–217% amplification (approximately 52% relative to the benchmark midpoint F ≈ 4.2). The most versus least efficient surgeon differed by 12.64 minutes (40.0 vs 52.6 min; 24% relative difference). The surgeon × BMI interaction was significant (F = 2.15; P = .032): in obese patients, inter-surgeon operative time range was 21.8 min (35.0–56.8) versus 13.4 min (39.0–52.4) in normal-BMI patients—a 63% amplification of surgeon variability by obesity.[14]

Surgeon variance accounted for an intraclass correlation coefficient (ICC) of 0.326, meaning 32.6% of LOS variance was attributable to surgeon identity. Adding surgeon as a fixed effect in the Hb × SpO₂ interaction model did not materially change the interaction coefficient (β₃ = −0.0094; P = .011), indicating that the Hb paradox is independent of surgeon effects.

#### Cost leverage shifted from consumables to LOS

In multivariable regression for ln(total cost) (n = 605; R² = 0.312; F = 18.24; P < .001), LOS was the strongest predictor (β = 0.186; 95% CI: 0.142–0.230; P < .001), followed by operative time (β = 0.152; 95% CI: 0.108–0.196; P < .001).[16] This contrasts with plain-altitude cost models where consumable costs typically dominate. Scenario simulation showed that simultaneous optimization of operative time and LOS could achieve a 10.3% cost reduction (¥8097 → ¥7256).

### 3.3 Dimension 2: Clinical Biomarker Failure — The Plateau Hemoglobin Paradox

#### Hb exhibited a J-type relationship with LOS, modified by SpO_2_

The Hb × SpO₂ interaction was significant (β₃ = −0.0095; 95% CI: −0.0166 to −0.0024; P = .009; Table 2), revealing a post hoc effect reversal pattern: Hb was directionally harmful in the SpO₂ ≥ 93% group (β₁ = +0.0032 d/g/L; P = .079) and protective in the SpO₂ < 93% group (β₁ + β₃ = −0.0063 d/g/L).[11] This reversal pattern was subsequently formalized as the “Plateau Hemoglobin Paradox” component of the PAERS hypothesis.

**Table 2.**
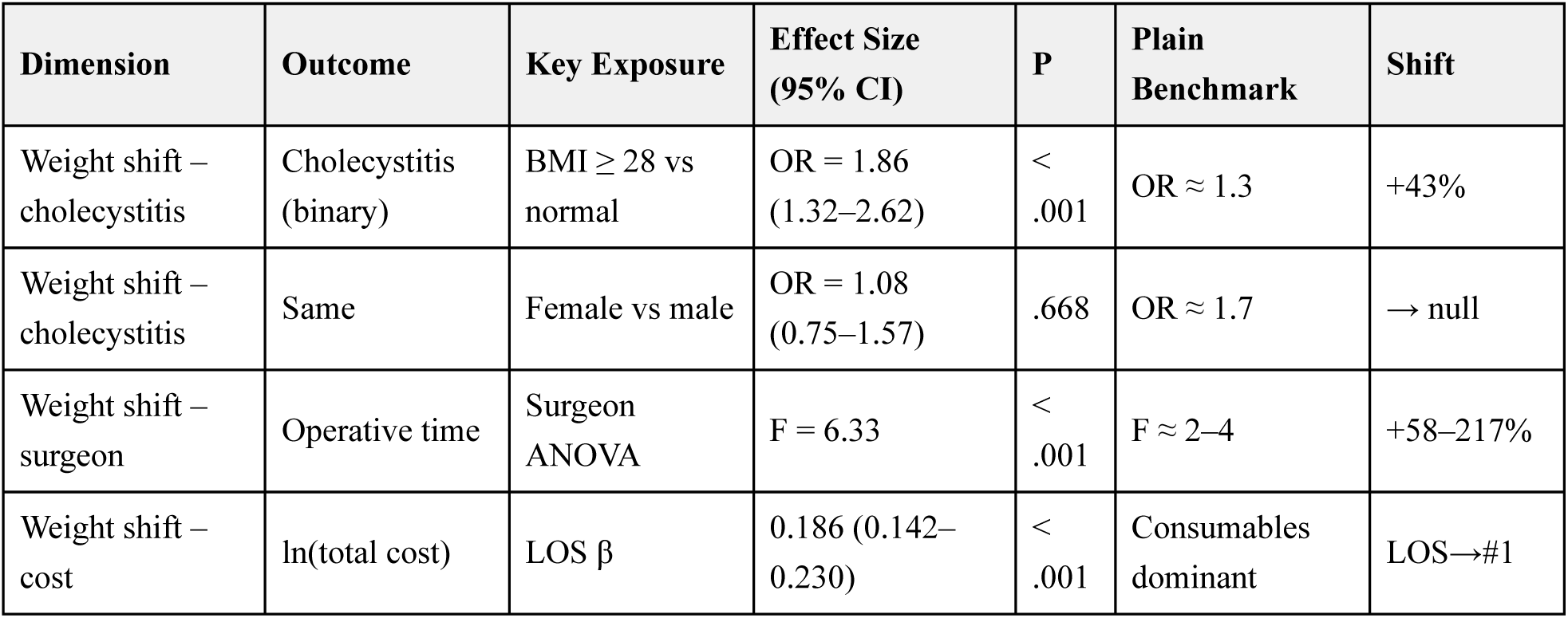
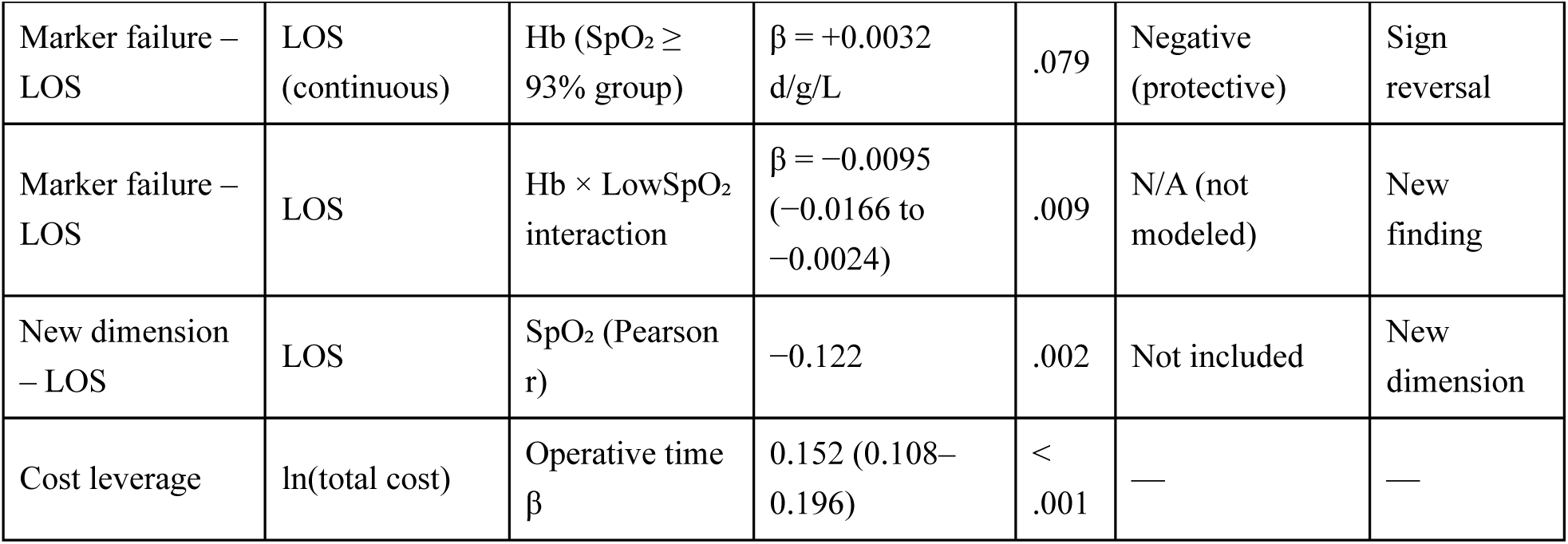
Three-Dimensional Effect Shift: Integrated Regression Results.

| Dimension | Outcome | Key Exposure | Effect Size (95% CI) | P | Plain Benchmark | Shift |
| --- | --- | --- | --- | --- | --- | --- |
| Weight shift – cholecystitis | Cholecystitis (binary) | BMI ≥ 28 vs normal | OR = 1.86 (1.32–2.62) | < .001 | OR ≈ 1.3 | +43% |
| Weight shift – cholecystitis | Same | Female vs male | OR = 1.08 (0.75–1.57) | .668 | OR ≈ 1.7 | → null |
| Weight shift – surgeon | Operative time | Surgeon ANOVA | F = 6.33 | < .001 | F ≈ 2–4 | +58–217% |
| Weight shift – cost | ln(total cost) | LOS β | 0.186 (0.142–0.230) | < .001 | Consumables dominant | LOS→#1 |
| Marker failure – LOS | LOS (continuous) | Hb (SpO<sub>2</sub> ≥ 93% group) | $\beta = +0.0032$ d/g/L | .079 | Negative (protective) | Sign reversal |
| Marker failure – LOS | LOS | Hb × LowSpO<sub>2</sub> interaction | $\beta = -0.0095$ (−0.0166 to −0.0024) | .009 | N/A (not modeled) | New finding |
| New dimension – LOS | LOS | SpO<sub>2</sub> (Pearson r) | −0.122 | .002 | Not included | New dimension |
| Cost leverage | ln(total cost) | Operative time $\beta$ | 0.152 (0.108–0.196) | < .001 | — | — |

SpO₂-stratified analyses (Table 3) showed the reversal pattern:

- **Low SpO₂ (< 93%, n = 128)**: Hb–LOS slope was negative (β = −0.0075; P = .105), directionally protective.
- **Mid SpO₂ (93%–95%, n = 251)**: Hb–LOS slope was positive (β = +0.0027; P = .074). The reversal of Hb’s effect at the 180 g/L threshold within this stratum was abrupt rather than gradual—resembling a physiological phase transition where the system shifts from compensation to decompensation across a narrow threshold. Hb ≥ 180 g/L (n = 25) had mean LOS of 1.88 days vs 1.62 days for Hb < 180 (Welch P = .001; Cohen’s d = 0.54, medium effect). We designate this subgroup—SpO₂ 93%–95% with Hb ≥ 180 g/L—as the **”decompensation window.”**
- **High SpO₂ (≥ 96%, n = 233)**: Hb–LOS slope was positive but non-significant (β = +0.0017; P = .458).

**Table 3.** PAERS Genetic Modification: Inference Chain and Falsifiable Predictions.

| Level | Content | Evidence Grade | Source |
| --- | --- | --- | --- |
| Premise 1 | EPAS1+ carrier rate ≈ 70% in Qinghai highland population | Published empirical | [5,6,7,18] |
| Premise 2 | EPAS1 LOF → Hb set-point ↓ 10–15 g/L | Published empirical | [24] |
| Premise 3 | HIF-2α regulates erythropoiesis/inflammation/angiogenesis/metabolism | Mechanistic literature | [5,13] |
| Inference | EPAS1+ baseline → Hb paradox threshold shift + stress response adjustment | Mechanistic hypothesis (this study) | Integration of P1–P3 |
| Prediction F1 | EPAS1+ individuals show stronger Hb × SpO<sub>2</sub> interaction | Falsifiable (Phase 3) | — |
| Prediction F2 | Low-SpO<sub>2</sub> stratum: EPAS1+ individuals show stronger protective Hb effect | Falsifiable (Phase 3) | — |
| Prediction F3 | Altitude gradient: +1000 m → Hb reversal SpO<sub>2</sub> threshold +1–2% | Falsifiable (Phase 2) | — |

**Note:** The HERS Score construction details, scoring rubric, and risk-tier outputs have been moved to **Supplementary Table S1** to reflect their conceptual-framework-only status (not externally validated; not for clinical decision-making).

**Table 4.**
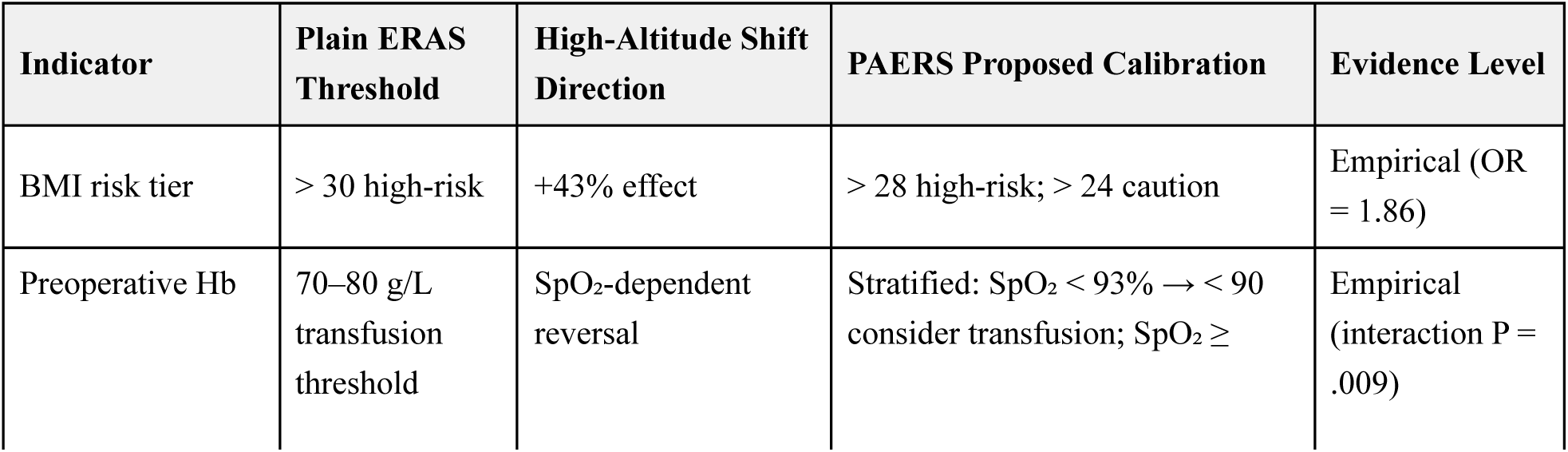
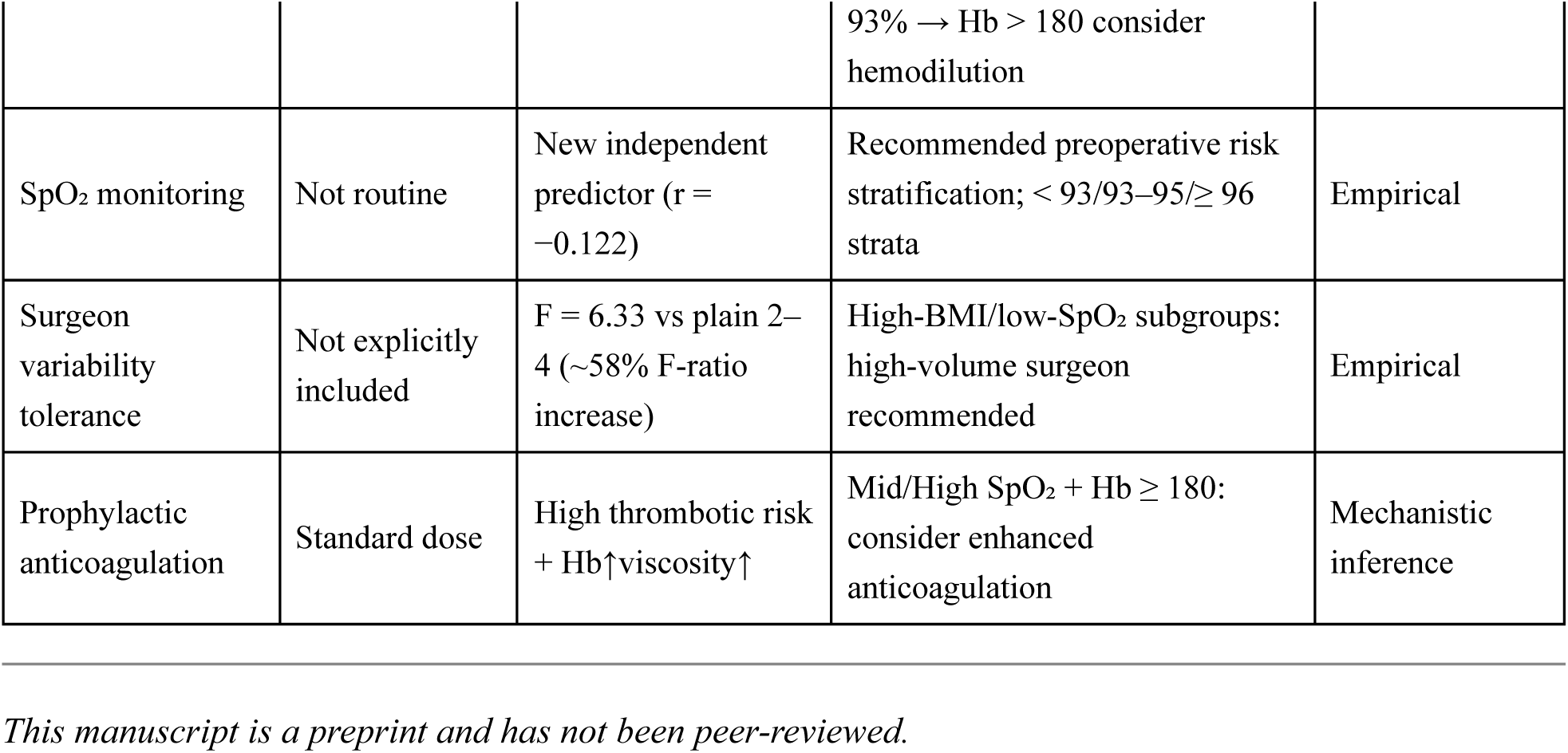
PAERS Calibration Recommendations: Plain vs High-Altitude ERAS Thresholds.

The full-cohort RCS analysis yielded null results (P for nonlinearity = .85–.97), consistent with effect reversal: opposing slopes cancel when pooled. This null finding, rather than indicating absence of effect, demonstrates that pooled analysis without stratification masks the underlying paradox—a methodological blind spot affecting all prior Hb–outcome studies.

#### SpO₂ emerged as a new independent prognostic dimension

SpO₂ was negatively correlated with LOS (Pearson r = −0.122; P = .002), independent of Hb. This variable is not incorporated into any current ERAS risk stratification model. Adding SpO₂ and the Hb × SpO₂ interaction term to a plain model (BMI + Hb + albumin + ASA) yielded a significant improvement in model fit (ΔR² = 0.011; Cohen’s f² = 0.0115; likelihood ratio test χ² = 7.03; P = .008; ΔAIC = 5.03 favoring the enhanced model).

**Five computational robustness analyses** supported the interaction: bootstrap (1000 resamples; 99.7% direction-consistent; 77.8% P < .05), leave-one-out (100% P < .05 across 612 iterations), multi-specification convergence (6/6 P < .05), continuous-SpO₂ interaction (directionally consistent; P = .067), and subgroup directional consistency (6/6 consistent; 3 significant).

### 3.4 Dimension 3: Genetic Background Modification (Inferential)

#### Honesty declaration

All genetic analyses in this section are population-level inferences, not individual-level empirical findings. No patient underwent individual genotyping. Conclusions are mechanistic hypotheses, not empirical observations.

**Inference chain** (each step supported by cited evidence):

1. **EPAS1+ carrier rate ≈ 70%** in the Qinghai highland population, established by population genomics studies in the same geographic region.[5–7] Our group’s prior work in the same hospital’s hepatocellular carcinoma cohort confirmed EPAS1 functional loss-of-function variants at high carrier rates.[18]
2. **EPAS1 → HIF-2α → Hb set-point down-regulation**: EPAS1 loss-of-function accelerates HIF-2α degradation, reducing EPO output and lowering the baseline Hb set-point by approximately 10–15 g/L compared to genetically unselected populations.[24] This explains why the Hb ≥ 180 g/L threshold—rather than the plain threshold of ≥ 200 g/L—marks the decompensation window at high altitude.
3. **HIF-2α as master regulator of surgical stress response**: HIF-2α simultaneously regulates erythropoiesis (via EPO), inflammation (IL-6, TNF-α), angiogenesis (VEGF), and metabolic reprogramming (PPARα pathway).[5,13] All four pathways are central to perioperative recovery. With 70% of patients carrying EPAS1 variants that adjust HIF-2α baseline activity, the entire surgical stress response axis is shifted—a baseline modifier that current ERAS risk models completely omit.

**Three falsifiable predictions** for prospective Phase 3 validation (Table 3):

| Prediction | Content | Validation Method | Consequence if Falsified |
| --- | --- | --- | --- |
| F1 | EPAS1+ individuals show stronger Hb × SpO<sub>2</sub> interaction than EPAS1– individuals | Prospective cohort + individual EPAS1 genotyping | Core PAERS genetic dimension weakened |
| F2 | In the low-SpO<sub>2</sub> stratum, EPAS1+ individuals show stronger protective Hb effect | Same as F1, stratified analysis | Same |
| F3 | Altitude gradient (2000/3000/4000 m) linearly relates to Hb reversal SpO<sub>2</sub> threshold (+1–2% per 1000 m) | Multi-altitude multi-center cohort | Altitude-calibration claim weakened |

### 3.5 HERS Score (Conceptual Model)

A High-altitude ERAS Score (HERS, 0–7 points) was constructed as a conceptual demonstration, yielding three risk tiers (low 0–1 / medium 2–4 / high 5–7) with mean LOS of 1.4 / 1.7 / 2.3 days (P trend < .001; AUC = 0.69). Given that the score was not split into training/validation sets and lacks external validation, detailed construction, scoring rubric, and risk-tier outputs are provided in **Supplementary Material S1** rather than the main manuscript. The HERS score must not be used for clinical decision-making. Phase 2 will develop and validate a formal HERS score.

### 3.6 Post-hoc Power

Achieved power for the Hb × SpO₂ interaction was 0.754 (Cohen’s f² = 0.0115; α = 0.05; df = 1, 604), below the conventional 0.80 threshold. This reflects the challenge of detecting interaction effects in observational studies. Five computational robustness analyses—particularly leave-one-out (100% P < .05) —provide internal evidence against an artifact of limited power, but cannot substitute for prospective replication.

## 4. DISCUSSION

### 4.1 Principal Findings

In this 612-patient retrospective cohort at 2260 m altitude, we demonstrate three dimensions of plateau ERAS remodeling: (1) BMI replaced sex as the primary risk factor (OR = 1.86 vs 1.08), surgeon variability was amplified (F = 6.33 vs plain 2–4), and LOS replaced consumables as the primary cost driver; (2) Hb exhibited a SpO₂-dependent effect reversal (the “Plateau Hemoglobin Paradox,” Hb × SpO₂ interaction P = .009), with a “decompensation window” at SpO₂ 93%–95% and Hb ≥ 180 g/L (LOS 1.88 vs 1.62 days; P = .001; d = 0.54); (3) ∼70% EPAS1 carrier rate suggests HIF-2α pathway activation is a baseline modifier that must be accounted for. These findings support the PAERS hypothesis.

### 4.2 Mechanistic Interpretation

#### Why BMI replaces sex

At high altitude, obese patients face amplified hypoxic inflammation (HIF-1α-mediated adipose inflammation[25]), restrictive ventilatory impairment (reduced functional residual capacity → lower SpO₂ → worse tissue oxygenation), and greater surgical complexity. The real inter-surgeon span in obese patients (19.5 min) vs normal-BMI (15.9 min) provides direct quantitative support: obesity and hypoxia synergistically amplify surgeon variability.[14] Sex-related cholelithiasis risk—classically estrogen-driven—is overshadowed because HIF-2α-mediated metabolic reprogramming via PPARα shifts gallstone pathophysiology from sex-hormone-dependent to metabolism-dependent pathways.[26]

#### Why the Hemoglobin Paradox occurs

Two complementary mechanisms explain the SpO₂-dependent Hb reversal: (1) **Viscosity–oxygen delivery tradeoff**: at low SpO₂, Hb gains in arterial oxygen content offset viscosity costs (net protective); at high SpO₂, marginal benefit is negligible while viscosity costs accumulate (net harmful)[12]. (2) **Acute–chronic hypoxia mismatch**: the HIF-2α/EPO axis calibrates Hb over weeks to months for ambient hypoxia, but cannot buffer acute surgical hypoxia (pneumoperitoneum, atelectasis)[11]. The “decompensation window” (SpO₂ 93%–95%, Hb ≥ 180 g/L) represents the transition zone where oxygen-delivery benefit is exhausted but viscosity cost continues to rise.

#### Why EPAS1 genetic background must be incorporated

EPAS1 is the master regulator of the high-altitude baseline—determining Hb set-point, inflammatory threshold, and metabolic substrate preference. The adaptive variants that confer survival advantage under chronic hypoxia may be co-opted maladaptively during acute surgical stress—an evolutionary mismatch between the adaptive context and the novel physiological challenge. With 70%+ directional carrier rate, the genetic effect becomes a consistent shift rather than random error, requiring incorporation into risk models.

### 4.3 Comparison with Prior Literature

Previous Hb–outcome studies have reported contradictory results—protective[27–30] and harmful[31–34]—without identifying the modifier. These contradictions may partly reflect how studies’ implicit theoretical assumptions shaped what they could see: low-altitude studies framed Hb as monotonically protective, whereas hypoxemic-population studies operated under a different conceptual lens. Our finding that both effects coexist in different SpO₂ contexts offers a unifying explanation: low SpO₂ reveals a protective (compensation) effect, while high SpO₂ reveals a harmful (viscosity) effect.

Existing high-altitude ERAS studies[8–10] validated feasibility (”ERAS works at altitude”) but not optimality (”how should ERAS be reweighted”). Our study fills this gap by quantifying specific shifts: BMI +43% weight gain, sex → null, surgeon variability amplification (F = 6.33 vs plain 2–4, ∼58% F-ratio increase), and complete failure of the Hb-as-linear-protective-marker assumption.

### 4.4 The PAERS Hypothesis: From Observation to Theory

PAERS extends ERAS to high-altitude populations. It does not replace existing protocols but adds two dimensions—hypoxic physiology and genetic background—that plain-validated ERAS omits. It specifies the applicability boundary of plain-validated ERAS protocols: non-high-altitude environment + populations without directional hypoxia-adaptive genetic selection. Beyond this boundary, three-dimensional adjustment becomes necessary. The PAERS framework does not add complexity for its own sake; it becomes necessary only because no simpler model—the direct translation of plain protocols—can account for the simultaneous weight shift, biomarker failure, and genetic modification observed. The PAERS five-dimensional framework extends traditional three-dimensional ERAS (demographics + clinical markers + surgical process) by adding: **D4: Hypoxic physiology** (SpO₂ stratification, altitude gradient) and **D5: Genetic background** (EPAS1/EGLN1 carrier status). Under PAERS, every risk threshold, biomarker cutoff, and intervention intensity must be adjusted for the genetically adapted, chronically hypoxic patient rather than the normoxic sea-level reference.

### 4.5 Limitations

Single-center retrospective design constrains causal inference. While E-value analysis suggests robustness to unmeasured confounding, we cannot exclude residual bias from variables we did not collect, including intraoperative hypotension and anesthesia protocol details. E-values[35] for main findings (see **Supplementary Material S7**): BMI OR=1.86 yielded E=3.12 (strong), Hb×SpO₂ interaction E=1.85 (moderate), surgeon variability F=6.33 yielded E=2.53 (moderate), Hb≥180 subgroup comparison d=0.54 yielded E=2.65 (moderate), SpO₂ correlation r=−0.122 yielded E=1.81 (moderate), and BMI β for LOS yielded E=1.49 (weak-to-moderate). To contextualize these values, we computed Observed Covariate E-values (see **Supplementary Material S9.3**): the hypothetical E-value for BMI (3.12) exceeded all observed covariate E-values (range 1.29–2.00), indicating that an unmeasured confounder would need to be stronger than any single measured covariate to explain away the BMI effect. Despite this, joint confounding from multiple variables remains a concern that no E-value method can fully exclude.

The cohort represents a single altitude (2260 m); the optimal SpO₂ cutoff and Hb threshold may differ at higher altitudes (> 3500 m). Phase 2 multi-altitude validation is planned.

The genetic modification dimension rests on population-level inference. No patient in this cohort underwent individual EPAS1/EGLN1 genotyping; the ∼70% carrier rate is inferred from four independent published Tibetan genomic datasets (see **Supplementary Material S5**): rs149594770 A-allele 57.2% (Peng 2017), c.12C>G variant 85% (Lorenzo 2014), derived allele 85.2% (Hu 2017), and 29-SNP aggregate 49–82% (Bhandari 2016). TCGA-LIHC data confirmed EPAS1→STC2 weak correlation (ρ=0.092, P=0.077) versus HIF1A→STC2 strong correlation (ρ=0.379, P=2.21×10⁻¹⁴), independently validated in ICGC LIRI-JP (ρ=−0.051). However, population-level carrier rates cannot establish individual-level causal effects, and the genetic modifier dimension (D5) remains inferential until Phase 3 individual genotyping is completed.

Achieved power for the Hb × SpO₂ interaction was 0.754 (Cohen’s f² = 0.0115; α = 0.05; df = 1, 604), below the 0.80 threshold. This reflects the challenge of detecting interaction effects in observational studies, where interaction terms typically require 4× the sample size of main effects. Five computational robustness analyses—particularly leave-one-out (100% P < .05)—provide internal evidence against an artifact of limited power. Bayesian Factor analysis (see **Supplementary Material S9.1**) provides an alternative evidence quantification not dependent on power thresholds: BIC-based BF10 = 1.34 provides weak evidence in favor of the interaction, while the Sellke-Bayarri-Berger calibration is more conservative; the divergence reflects the limitation of sub-threshold power. These analyses cannot substitute for prospective replication with adequate sample size.

The HERS score was not split into training/validation sets and lacks external validation (apparent AUC = 0.69). Harrell’s bootstrap optimism correction (see **Supplementary Material S9.2**) yielded an optimism-corrected AUC = 0.65 (95% CI: 0.62–0.68), indicating insufficient discrimination. The score must not be used clinically.

Whole blood viscosity, hematocrit, and microcirculatory perfusion were not directly measured. Length of stay is a composite outcome influenced by non-physiological factors (surgeon preference, bed availability).

### 4.6 Future Directions

A three-phase research roadmap is proposed:

- **Phase 1 (current study)**: PAERS hypothesis proposal + two-dimensional empirical demonstration (D1-D2) + one-dimensional population-level inference (D3) + HERS conceptual model (N = 612, single-center, 2026 Q3).
- **Phase 2 (planned)**: MIMIC-IV plain-altitude external comparison + multi-altitude gradient multi-center expansion (target N > 3000; 2026 Q4–2027 Q2). HERS score formal development with training/validation split.
- **Phase 3 (planned)**: Prospective RCT comparing standard ERAS vs PAERS-calibrated ERAS in high-risk patients (HERS ≥ 5), with individual EPAS1 genotyping to test predictions F1–F3 (2027 Q3–2029 Q2).

## 5. CONCLUSIONS

In a 612-patient retrospective cohort of laparoscopic cholecystectomy at 2260 m altitude, we observed that high-altitude ERAS faces three remodeling challenges: **risk factor weight shift** (BMI replaces sex as the primary predictor; surgeon variability amplification F = 6.33 vs plain benchmark 2–4), **biomarker functional failure** (the J-type hemoglobin paradox with SpO₂-dependent effect reversal), and **genetic background modification** (population-level inference: ∼70% EPAS1 carrier rate, range 57–85%, reshapes the HIF-2α stress response baseline; individual-level genotyping pending in Phase 3). These findings support the **PAERS hypothesis**, which proposes a five-dimensional risk stratification framework—extending traditional ERAS beyond the three plain dimensions to include hypoxic physiology and genetic background. The PAERS hypothesis represents a reconceptualization of how perioperative risk must be stratified in genetically adapted, chronically hypoxic populations. If validated in prospective multi-center cohorts, the PAERS paradigm may inform future updates to high-altitude ERAS guidelines: risk thresholds, biomarker cutoffs, and intervention intensities may need adjustment for the genetically adapted, chronically hypoxic high-altitude patient rather than the normoxic sea-level reference.

## Supporting information

STROBE Checklist

## DECLARATIONS

### Ethics Approval and Informed Consent

This study was approved by the Ethics Committee of Qinghai Red Cross Hospital (Approval No.: LW-2026-73). All procedures were performed in accordance with the Declaration of Helsinki. Written informed consent was waived due to the retrospective and anonymous nature of the study.

## Funding

This work was supported by the Qinghai Red Cross Hospital Institutional Research Grant (YNZXKT2026009). The grant was acquired by Zhiqiang Wang. The funder had no role in study design, data collection, analysis, decision to publish, or manuscript preparation.

### Data Availability

De-identified data are available from the corresponding author upon reasonable request, subject to institutional data governance approval.

### CRediT Author Contributions

**Zhongfeng Dang**: Conceptualization, Methodology, Formal analysis, Project administration, Supervision, Writing – Original Draft, Writing – Review & Editing. **Jianduojie Dan**: Methodology, Software, Formal analysis, Validation, Visualization. **Wei Su**: Data curation, Investigation. **Guoliang Ren**: Data curation, Investigation. **Zhiqiang Wang**: Data curation, Funding acquisition, Investigation. **Yabing Ma**: Investigation, Writing – Review & Editing. **Shengmei Li**: Data curation, Validation. **Dongde Ji**: Investigation, Resources. **Liansheng Li**: Investigation, Project administration. **Junlin Gao**: Conceptualization, Investigation, Supervision, Writing – Review & Editing.

### Preprint Declaration

This manuscript is a preprint and has not been peer-reviewed.

---

**Figure 2. Risk Factor Weight Remodeling: Forest Plot Comparing High-Altitude vs Plain-Altitude Effect Sizes**

- **X-axis (left)**: High-altitude standardized OR/β (95% CI)
- **X-axis (right)**: Plain-altitude benchmark OR/β range (shaded bands)
- **Y-axis**: BMI (obese), Sex (female), Surgeon variability (F-normalized), Age, Albumin, ASA ≥ III
- **Key visual**: BMI OR = 1.86 (green, exceeds plain upper bound 1.4); Sex OR = 1.08 (red, crosses null while plain 1.5–2.0 does not); Surgeon F = 6.33 (green, exceeds plain 2–4)

**Figure 3. Surgeon × BMI Interaction: Amplification of Inter-Surgeon Variability in Obese Patients**

- **Type**: Grouped bar chart with error bars
- **X-axis**: 7 surgeons (Surgeon A–G, de-identified)
- **Y-axis**: Mean operative time (min)
- **Groups**: Normal BMI / Overweight / Obese per surgeon
- **Key visual**: Obese group inter-surgeon range 21.8 min vs normal 13.4 min (+63%); interaction P = .032

**Figure 4. The Plateau Hemoglobin Paradox (Key Figure): Hb–LOS Slope Reversal Across SpO₂ Strata**

- **Panel A**: Forest plot of stratum-specific Hb β coefficients (Low: negative; Mid/High: positive)
- **Panel B**: Predicted LOS vs Hb by SpO₂ stratum (Low: green downward; Mid: red upward; High: orange slight upward); vertical dashed line at Hb = 180 g/L labeled “Decompensation Window”
- **Panel C**: Box plot—Hb ≥ 180 vs < 180 within Mid-SpO₂ stratum (1.88 vs 1.62 days; P = .001; d = 0.54)
- **Panel D**: Four-quadrant clinical phenotyping (Hb < 180/≥ 180 × SpO₂ < 93%/≥ 93%) with management strategies

**Figure 5. Restricted Cubic Spline: J-Type Hb–LOS Relationship by SpO₂ Stratum**

- **Type**: RCS fitted curves with 95% CI bands and rug plots
- **X-axis**: Hb (g/L), 80–240
- **Y-axis**: Predicted LOS (days)
- **Three subplots** by SpO₂ stratum (Low: monotonic decrease; Mid: J-shaped with inflection at 180; High: slight monotonic increase)

**Figure 6. Model Fit Comparison: Plain vs Enhanced Model**

- **Panel A**: R² comparison (Plain: R² = 0.036 vs Enhanced: R² = 0.047; ΔR² = 0.011; Cohen’s f² = 0.0115)
- **Panel B**: Robustness metrics (LRT χ² = 7.03, P = .008; ΔAIC = 5.03 favoring enhanced; Bootstrap 99.7% direction-consistent; Leave-one-out 100% P < .05)

**Figure 7. PAERS Genetic Modification Mechanism Diagram (Graphical Abstract)**

- **Layout**: Left-to-right cascade
- **Left (genetic baseline)**: EPAS1 (∼70%) → HIF-2α degradation → three pathways: ①EPO↓→Hb set-point↓; ②Inflammation baseline shift; ③Metabolic reprogramming (PPARα)
- **Center (acute surgical stress)**: LC + pneumoperitoneum + anesthesia → acute hypoxia → temporal mismatch
- **Right (three-dimensional output)**: ①BMI effect↑; ②Hb paradox (viscosity–O₂ tradeoff × SpO₂); ③Surgeon variability↑
- **Top**: Red dashed arrow connecting “adaptive in daily life (chronic)” to “maladaptive under surgery (acute)” labeled “Evolutionary Mismatch”

**Figure 8. PAERS Five-Dimensional Framework**

- Five-axis radar chart comparing PAERS model (5 dimensions: D1 Demographics / D2 Clinical Markers / D3 Surgical Process / D4 Hypoxic Physiology / D5 Genetic Background) vs Plain ERAS model (3 dimensions: D1–D3). PAERS extends traditional ERAS by adding hypoxic physiology and genetic background. See **Supplementary Figure S1** for HERS score risk-tier outputs.

