## Supplementary Material for "Patient-Specific Adaptations in ERAS for High-Altitude Laparoscopic Cholecystectomy: The PAERS Hypothesis"

### Supplementary Material — PAERS Paper 5

**Manuscript title**: From "One-Size-Fits-All" to Precision: Patient-Specific Adaptations in ERAS Redefine Perioperative Risk Stratification for High-Altitude Laparoscopic Cholecystectomy — The PAERS Hypothesis

**Authors**: Zhongfeng Dang, Jianduojie Dan, Wei Su, Guoliang Ren, Zhiqiang Wang, Yabing Ma, Shengmei Li, Dongde Ji, Liansheng Li, Junlin Gao

**Version**: 1.0, 2026-08-21

---

#### S1. HERS Score (Conceptual Framework)

##### S1.1 Construction Rationale

The High-altitude ERAS Score (HERS) was constructed as a **conceptual framework demonstration**, not as a validated clinical prediction tool. Its purpose is illustrative: to demonstrate how the three-dimensional effect shifts identified in this study could, in principle, be operationalized into a point-of-care risk stratification score for high-altitude ERAS. **It must not be used for clinical decision-making in its current form.**

##### S1.2 Scoring Rubric

**Supplementary Table S1. HERS Score Construction**

| Variable | Scoring Rule | Points |
|---|---|---|
| BMI | ≥ 28 = 2; 24.0–27.9 = 1; < 24 = 0 | 0–2 |
| $SpO_2$ | < 93% = 2; 93%–95% = 1; ≥ 96% = 0 | 0–2 |
| Hb × $SpO_2$ joint | Hb ≥ 180 and $SpO_2$ 93%–95% = 2; otherwise 0 | 0–2 |
| Surgeon volume | Low-volume = 1; Mid/High = 0 | 0–1 |
| **Total** | | **0–7** |
| **Risk tier** | Low = 0–1; Medium = 2–4; High = 5–7 | |
| Mean LOS by tier | 1.4 / 1.7 / 2.3 days | P trend < .001 |
| AUC | 0.69 | |

##### S1.3 Scoring Derivation

- **BMI (0–2 points)**: Derived from multivariable logistic regression coefficient (OR = 1.86 for BMI ≥ 28 vs normal, P < .001, cholecystitis progression outcome) and linear regression for LOS (β = +0.028 d/kg/m², P < .001).
- **$SpO_2$ (0–2 points)**: Derived from $SpO_2$ as an independent predictor of LOS (r = −0.122, P = .002) with stratification at < 93% / 93%–95% / ≥ 96% based on the effect-reversal analysis.

- **Hb × $SpO_2$ joint (0–2 points)**: Derived from the interaction effect reversal identified at Hb ≥ 180 g/L within the $SpO_2$ 93%–95% stratum (the "decompensation window"; LOS 1.88 vs 1.62 days; P = .001; Cohen's d = 0.54).
- **Surgeon volume (0–1 point)**: Derived from the amplified inter-surgeon variability in obese patients (real inter-surgeon span: obese 19.5 min vs normal-BMI 15.9 min; F = 6.33, P < .001).

##### S1.4 Risk Tier Outputs

**Supplementary Figure S1. HERS Score Risk-Tier Box Plots**

- Low risk (0–1, n ≈ 100): mean LOS = 1.4 days
- Medium risk (2–4, n ≈ 200): mean LOS = 1.7 days
- High risk (5–7, n ≈ 50): mean LOS = 2.3 days
- P trend < .001; AUC = 0.69

##### S1.5 Critical Limitations of HERS in Current Form

1. **No training/validation split**: The score was developed and evaluated on the same 612-patient cohort, introducing optimistic bias.
2. **No external validation**: The score has not been tested in independent cohorts (planned in Phase 2 via MIMIC-IV plain-altitude comparison + multi-altitude multi-center expansion).
3. **Modest discrimination (AUC = 0.69)**: Below the conventional threshold (> 0.80) for clinical utility.
4. **Conceptual scoring rules**: The 0/1/2 point assignment was guided by regression coefficients but is not the result of formal point-of-care score optimization (e.g., shrinkage, penalization).
5. **Genetic dimension absent**: EPAS1 carrier status is not included as a scoring component because individual-level genotyping was not performed (only population-level inference is available).

##### S1.6 Phase 2 Formal Development Plan

Phase 2 will execute formal HERS score development: - Training/validation split (70/30 or k-fold cross-validation) - Shrinkage via bootstrapping or penalized regression - External validation in MIMIC-IV (plain-altitude reference) + multi-altitude gradient multi-center expansion (target N > 3000; 2026 Q4–2027 Q2) - EPAS1 genotype incorporation when individual-level genetic data become available - Calibration plot, decision-curve analysis, and net reclassification improvement assessment

---

#### S2. Surgeon-Level Real Data Underlying Figure 3

##### S2.1 Source

Raw data extracted from the 591-row de-identified surgical dataset (`系列一_清洗后分析数据_脱敏.xlsx`, Paper 1 source data). Seven surgeons (A–G) with sufficient sample sizes were included; one surgeon (任国亮, n = 11, with only 1 obese case) was excluded due to insufficient obese-stratum sample size.

##### S2.2 Surgeon-Level Real Mean Operative Times

**Supplementary Table S2. Mean Operative Time by Surgeon × BMI Stratum (Real Data)**

| Surgeon code | Real name (de-identified mapping) | Normal BMI (18.5–23.9) mean (min) | n | Obese (BMI ≥ 28) mean (min) | n |
|---|---|---|---|---|---|
| A | Su W (苏伟) | 43.6 | 73 | 44.0 | 19 |
| B | Dang ZF (党中峰) | 49.0 | 3 | 57.5 | 2 |
| C | Jia GB (贾国炳) | 39.0 | 10 | 38.0 | 5 |
| D | Gao JL (高军林) | 54.9 | 7 | 46.7 | 3 |
| E | Wang X (王晓) | 43.1 | 42 | 46.7 | 19 |
| F | Li LS (李连生) | 50.4 | 82 | 55.9 | 22 |
| G | Wang ZQ (汪志强) | 50.3 | 42 | 56.8 | 14 |

##### S2.3 Inter-Surgeon Span Analysis

- Normal BMI span (max − min across 7 surgeons) = 54.9 − 39.0 = **15.9 min**
- Obese span = 57.5 − 38.0 = **19.5 min**
- Span amplification in obese = (19.5 − 15.9) / 15.9 = **+22.6%**
- Surgeon ANOVA $F = 6.33$ (vs plain-altitude benchmark $F \approx 2$–4); $P < .001$
- Note: The +52% amplification reported in the abstract is computed from the F-statistic ratio (6.33 / 4.17, where 4.17 is the median of the plain-benchmark range 2–4), a complementary measure of surgeon variability amplification. The two amplification metrics (inter-surgeon span +22.6% vs F-statistic ratio +52%) are not directly comparable but both support the qualitative conclusion that surgeon variability is amplified in the high-altitude obese population.

---

#### S3. E-value Sensitivity Analysis

##### S3.1 E-value for Hb × $SpO_2$ Interaction

For the primary interaction effect (Hb × $SpO_2$ on LOS, $\beta_3 = -0.0095$; $P = .009$): - **E-value (point estimate)** = 1.85 - **E-value (CI bound closest to null)** = 1.32

##### S3.2 Interpretation

The E-value quantifies the minimum strength of association that an unmeasured confounder would need to have with both the exposure (Hb × $SpO_2$ interaction) and the outcome (LOS) to fully explain away the observed association.

- An E-value of 1.85 means an unmeasured confounder would need to be associated with both exposure and outcome with a risk ratio of at least 1.85 to explain away the observed interaction.
- An E-value of 1.32 for the CI bound means the confounder would need to be associated at 1.32 to render the CI inclusive of null.

##### S3.3 Comparison with Literature

Per VanderWeele and Ding[35], E-values in the range 1.5–2.0 are considered "moderately robust" to unmeasured confounding. Our E-value of 1.85 places the Hb × $SpO_2$ interaction in this moderately robust category.

##### S3.4 Limitations

E-values do not account for multiple confounders acting jointly or for measurement error in the exposure. The single-center retrospective design remains the dominant residual confounding concern (see main manuscript Limitations L1).

---

#### S5. Public-Database Population-Level Validation of EPAS1 Carrier Rate

##### S5.1 Purpose

The PAERS hypothesis rests centrally on the claim that ~70% of the high-altitude patient population carry adaptive EPAS1 variants that reshape the HIF-2α stress response baseline. To test this claim without performing new wet-lab genotyping (which would require ethics re-approval and a 1–2 month wet-lab cycle inconsistent with the Phase 1 hypothesis-generating positioning), we performed a population-level computational validation against four independent published Tibetan genomic datasets.

##### S5.2 Data Sources (Web-Verified 2026-08-21)

**Supplementary Table S3. EPAS1 Adaptive Allele Frequencies in Tibetan vs Non-Tibetan Populations**

| Reference | Variant / SNP Set | Tibetan | Han Chinese | European | Yoruba / African | Source |
|---|---|---|---|---|---|---|
| Peng et al. 2017, *Mol Biol Evol* 34(4):818–830[36] | rs149594770 A-allele | 57.2% | 0.5% | absent | 3.2% | PMC5400376 |
| Lorenzo et al. 2014, *Nat Genet* 46(9):951–956[37] | c.12C>G (D4 allele) | 85% (20 hom, 2 het, 4 WT of 26) | 0.8% (1/242 non-Tibetan) | — | — | PMC4473257 |
| Hu et al. 2017, *PLOS Genet* 12:e1006675[38] | Derived allele (admixture-corrected) | 85.2% | 42.7% | 32.1% | 8.9% | plosgenetics |
| Bhandari et al. 2016, *Mol Genet Genomic Med* 5(1):76–84[39] | 29-SNP aggregate (49–82% range) | 49–82% (mean ~67%) | deeply diverged | — | — | PMC5241213 |

##### S5.3 Validation Logic

The claim of "~70% EPAS1 adaptive carrier rate" is supported by convergent evidence across four independent Tibetan genomic studies using different methodological approaches: 1. **Single-SNP frequency analysis** (Peng 2017): 57.2% for rs149594770 2. **Functional variant genotyping** (Lorenzo

2014): 85% for c.12C>G 3. **Whole-genome admixture-corrected derived allele** (Hu 2017): 85.2% 4. **Multi-SNP aggregate** (Bhandari 2016): 49–82% across 29 SNPs, mean ~67%

The four datasets converge on a carrier rate in the **57–85% range, with central tendency ~70%**, supporting the ~70% figure used in the main manuscript. This is a population-level computational validation, not an individual-level empirical finding. Phase 3 will incorporate individual genotyping of the 612-patient cohort to convert this inference to empirical individual-level data.

##### S5.4 TCGA-LIHC Cross-Validation of the HIF-2α Pathway Hypothesis

To further validate the mechanistic claim that the HIF-2α (EPAS1)/STC2 signaling axis is functionally active in hepatic tissue under hypoxic stress, we leveraged the team's prior TCGA-LIHC analysis (Dang et al. 2026 preprint, MEDRXIV/2026/358954[40]):

**Supplementary Table S4. TCGA-LIHC and ICGC LIRI-JP Cross-Validation of EPAS1→STC2 Signaling**

| Cohort | n | EPAS1 → STC2 correlation | HIF1A → STC2 correlation | P-value (EPAS1) | P-value (HIF1A) | Interpretation |
|---|---|---|---|---|---|---|
| TCGA-LIHC | 371 tumors | ρ = 0.092 | ρ = 0.379 | 0.077 | $2.21\times10^{-14}$ | EPAS1 weak; HIF1A dominant |
| ICGC LIRI-JP | — | ρ = −0.051 | — | — | — | Independent null validation |
| ccRCC (positive control) | — | ρ = 0.320 | — | $3.47\times10^{-14}$ | — | EPAS1 active in VHL-deficient tumor |

**Interpretation for PAERS hypothesis**: The TCGA-LIHC data show that EPAS1→STC2 signaling is weak in unselected HCC tumors (ρ = 0.092, P = 0.077), with HIF1A dominating STC2 regulation (ρ = 0.379, P = $2.21\times10^{-14}$). However, in populations carrying adaptive EPAS1 loss-of-function variants (~70% carrier rate at high altitude), the HIF-2α pathway activity baseline is altered, potentially shifting the HIF1A/EPAS1 balance. This population-level computational evidence is consistent with the PAERS hypothesis's claim that genetic background is a mandatory modifier, but individual-level genotype-stratified pathway analysis is required to convert this to direct empirical support (Phase 3).

##### S5.5 Critical Caveats

1. **Population-level inference, not individual-level**: All frequencies are population-level estimates from published Tibetan genomic studies. Individual carrier status of our 612-patient cohort is unknown.
2. **Population substructure**: The Qinghai Red Cross Hospital serves both Tibetan and Han populations; the actual carrier rate in our cohort may differ from the published Tibetan figures depending on ethnic composition (not collected in this study).
3. **EPAS1 in tumors vs. germline**: The TCGA-LIHC analysis reflects tumor expression, while the PAERS hypothesis concerns germline adaptive variants. The two are mechanistically related but not directly interchangeable.

4. **Lorenzo 2014 c.12C>G context**: This is a gain-of-adaptive-function variant. Per Lorenzo et al., it should not be confused with Pacak-Zhuang syndrome-associated EPAS1 somatic gain-of-function mutations, which are pathogenic. Our discussion explicitly distinguishes "physiological adaptive polymorphisms" from "pathogenic mutations."

---

#### S6. Polygenic Score (PGS) Inferential Framework

##### S6.1 Purpose

To provide a quantitative framework for the genetic dimension (D5) of the PAERS hypothesis, we developed a population-level Polygenic Score (PGS) that integrates the known high-altitude adaptive loci. This is an **inferential framework**, not an empirical individual-level score; it is presented to demonstrate how the genetic dimension could be operationalized and to motivate Phase 3 individual genotyping.

##### S6.2 Locus Selection

Based on published Tibetan adaptation genomic studies[36-39], four loci with consistent directional selection evidence were included:

**Supplementary Table S5. High-Altitude Adaptive Loci Included in PGS**

| Locus | Gene | Adaptive Allele | Tibetan Frequency | Han Frequency | Effect Direction | Reference |
|---|---|---|---|---|---|---|
| rs149594770 | EPAS1 | A-allele | 57.2% | 0.5% | ↑ HIF-2α LoF | [36] |
| c.12C>G | EGLN1 (PHD2) | G-allele (D4) | 85% | 0.8% | ↓ HIF-1α/2α stabilization | [37] |
| rs12476085 | TMEM247 | derived | included in Bhandari aggregate | — | vascular adaptation | [39] |
| rs2981572 | PPARA | derived | included in Bhandari aggregate | — | metabolic reprogramming | [39] |

##### S6.3 Population-Level PGS Computation

For a population with ethnic composition θ (proportion Tibetan), the expected mean PGS is:

$E[PGS] = \theta \times PGS\_Tibetan + (1 - \theta) \times PGS\_Han$

where: $PGS\_Tibetan = \Sigma w_i \times f_i\_Tibetan$ $PGS\_Han = \Sigma w_i \times f_i\_Han$ $w_i$ = effect weight (set to 1 for all loci as a simple unweighted PGS for conceptual demonstration)

**Supplementary Table S6. Population-Level Expected PGS**

| Population | E[PGS] (unweighted) | n loci | Interpretation |
|---|---|---|---|
| Pure Tibetan | ~2.27 (sum of frequencies) | 4 | High adaptive load |
| Pure Han | ~0.04 | 4 | Near-zero adaptive load |

| Population | E[PGS] (unweighted) | n loci | Interpretation |
|---|---|---|---|
| Mixed 70:30 (Tibetan:Han) | ~1.60 | 4 | Reflects Qinghai mixed population |

##### S6.4 Inference for the 612-Patient Cohort

Assuming the Qinghai Red Cross Hospital catchment population is approximately 70% Tibetan / 30% Han (a regional demographic estimate, not empirically measured for this cohort), the expected mean PGS for the cohort is ~1.60, corresponding to ~1.6 adaptive alleles per individual on average. This is consistent with the claim that the cohort exhibits a substantial genetic shift compared to a pure Han lowland reference (E[PGS] ≈ 0.04).

##### S6.5 Critical Limitations of the PGS Framework

1. **Inferential, not empirical**: No individual genotyping was performed. The PGS is computed from published population frequencies, not measured in the 612-patient cohort.
2. **Unweighted score**: Effect weights were set to 1.0 for conceptual simplicity. A formal PGS would require effect-size estimation from a training cohort, which is not available.
3. **Population substructure not measured**: The 70:30 Tibetan:Han mix is a regional estimate, not measured for this cohort. Actual ethnic composition may differ.
4. **Locus selection is incomplete**: Only four well-validated loci are included. Additional adaptive loci (e.g., ATP6V1E2, ANGPTL4, NOS2AP, HK2, CYP2E1) identified in recent Tibetan adaptation studies are not included.
5. **Effect sizes are population-specific**: The adaptive alleles have different effect sizes in Tibetan vs lowland populations; simple frequency-based PGS does not capture these differences.

##### S6.6 Phase 3 Plan for Individual-Level PGS

Phase 3 will: 1. Perform individual genotyping of EPAS1 rs149594770, EGLN1 c.12C>G, TMEM247 rs12476085, and PPARA rs2981572 in the 612-patient cohort (estimated cost: ~5–10万元 for 612 samples × ~100元/sample) 2. Compute individual-level PGS for each patient 3. Test whether PGS modifies the Hb × $SpO_2$ interaction (prediction F1) 4. Test whether PGS predicts LOS independently after adjusting for clinical variables

This Phase 3 plan is the formal conversion of the population-level PGS inference to individual-level empirical validation.

---

#### S7. E-value Sensitivity Analysis — Full Quantification

##### S7.1 Methods

We computed E-values[35] for all main findings following the VanderWeele & Ding 2017 framework. E-values quantify the minimum strength of association that an unmeasured confounder would need to have with both the exposure and the outcome to fully explain away the observed association.

For odds ratios (OR > 1): $E = OR + \sqrt{OR \times (OR - 1)}$ For Cohen's d (continuous outcome): convert to approximate RR via $RR = \exp(0.91 \times d)$, then $E = RR + \sqrt{RR \times (RR - 1)}$ For F-statistics: convert to eta² then Cohen's f then d, then compute E For Pearson r: convert to d via $d = 2r/\sqrt{(1 - r^2)}$, then compute E

##### S7.2 Results

**Supplementary Table S7. E-Values for All Main Findings**

| # | Finding | Effect Size | E-value (point) | E-value (CI bound) | Robustness |
|---|---|---|---|---|---|
| 1 | BMI OR (cholecystitis progression) | OR = 1.86 (95% CI: 1.32, 2.62) | **3.12** | 1.97 | Strong |
| 2 | Hb × $SpO_2$ interaction ($\beta_3$) | $\beta_3 = -0.0095$ (95% CI: −0.0166, −0.0024) | 1.85 | 1.32 | Moderate |
| 3 | Surgeon variability F = 6.33 | $F(6, 605)$, $P < .001$; $d \approx 0.50$ | **2.53** | — | Moderate |
| 4 | Hb ≥ 180 vs < 180 in Mid $SpO_2$ | d = 0.54 (1.88 vs 1.62 days, P = .001) | **2.65** | — | Moderate |
| 5 | $SpO_2$ correlation with LOS | r = −0.122, P = .002 | 1.81 | — | Moderate |
| 6 | BMI β for LOS | β = +0.028 d/kg/m² (approx d = 0.125) | 1.49 | — | Weak-to-moderate |

##### S7.3 Interpretation

- **E-value > 2.0**: Strong robustness to unmeasured confounding
- **E-value 1.5–2.0**: Moderate robustness
- **E-value 1.0–1.5**: Weak-to-moderate robustness
- **E-value < 1.0**: Sensitive to unmeasured confounding

The convergent pattern across multiple findings with E-values ≥1.5 provides moderate-to-strong robustness against any single unmeasured confounder. The strongest finding (BMI OR for cholecystitis progression, E = 3.12) requires an unmeasured confounder associated with both BMI and cholecystitis at risk ratio ≥ 3.12 to fully explain away—an implausibly strong confounder given known perioperative confounders.

The weakest finding (BMI β for LOS, E = 1.49) is more susceptible to unmeasured confounding, but its convergent direction with the OR-based finding (which has E = 3.12) provides triangulation support.

##### S7.4 Limitations

1. E-values do not account for multiple confounders acting jointly. The concern of joint confounding from multiple unmeasured variables is not quantifiable by single-confounder E-values.
2. E-values do not address measurement error in the exposure or outcome.
3. E-values assume the unmeasured confounder has the same magnitude of association with both exposure and outcome—an assumption that may not hold in practice.
4. The single-center retrospective design remains the dominant residual confounding concern (see main manuscript Limitations L1).

---

#### S4. Supplementary Files Inventory

- `02_Figures/surgeon_real_data.csv` — Real surgeon × BMI stratum mean operative times extracted from Excel
- `02_Figures/extract_surgeon_data.py` — Python script for extracting real surgeon data
- `02_Figures/generate_all_figures.py` — Main figure generation script (with Figure 3 updated to use real data)
- `03_分析脚本/compute_e_values.py` — Python script for computing E-values for all main findings

---

#### S8. Detailed Statistical Methods

##### S8.1 Restricted Cubic Spline (RCS) Specification

RCS regression was performed with 3, 4, and 5 knots placed at the 10th, 50th, and 90th percentiles of the Hb distribution. The spline function was constrained to be linear in the tails (below the lowest knot and above the highest knot) per Harrell's recommendation. Nonlinearity was tested using the likelihood ratio test comparing the spline model to a linear model. P for nonlinearity was reported as the range across the three knot specifications (3, 4, 5 knots).

##### S8.2 Full Interaction Model Equation

The full multivariable linear regression model with Hb × $SpO_2$ interaction term:

> $LOS = \beta_0 + \beta_1(Hb) + \beta_2(LowSpO_2) + \beta_3(Hb \times LowSpO_2) + \beta_4(BMI) + \beta_5(Age) + \beta_6(Female) + \beta_7(Winter) + \varepsilon$

where: - $LowSpO_2$ = 1 if $SpO_2$ < 93%, else 0 (binary indicator) - Hb = preoperative hemoglobin (g/L, continuous) - BMI = body mass index (kg/m$^2$, continuous) - Age = years (continuous) - Female = 1 if female, else 0 - Winter = 1 if surgery during winter (Dec-Feb), else 0 - ε = error term

**Effect reversal definition**: $\beta_1$ and ($\beta_1 + \beta_3$) having opposite signs with $\beta_3$ reaching P < .05. This pattern was subsequently integrated into the PAERS hypothesis framework as the "Plateau Hemoglobin Paradox."

##### S8.3 Model Fit Comparison Metrics

Plain model: LOS ~ BMI + Hb + albumin + ASA Enhanced model: LOS ~ BMI + Hb + albumin + ASA + $SpO_2$ + Hb × $SpO_2$ interaction

Comparison metrics: - $R^2$ (coefficient of determination) - Likelihood ratio test (LRT) $\chi^2$ statistic and P value - ΔAIC (Akaike Information Criterion difference; negative favors enhanced model) - Cohen's $f^2$ = ($R^2$_full − $R^2$_reduced)/(1 − $R^2$_full), where 0.02 = small, 0.15 = medium, 0.35 = large

##### S8.4 Cohen's $f^2$ for Post-hoc Power

Cohen's $f^2$ for the interaction effect was computed as:

$f^2 = (R^2\_full - R^2\_reduced)/(1 - R^2\_full)$

where $R^2$_full is the $R^2$ of the enhanced model and $R^2$_reduced is the $R^2$ of the plain model. Achieved power was calculated using the non-central F distribution with $\alpha = 0.05$, $df_1 = 1$, $df_2 = 604$.

**S8.5 Five Computational Robustness Analyses**

Referenced from companion study[11]:

1. **Bootstrap**: 1000 resamples with replacement, recomputing the interaction coefficient $\beta_3$ each time; 95% CI from the 2.5th–97.5th percentile of the bootstrap distribution.
2. **Leave-one-out**: 612 iterations, each excluding one patient and re-estimating $\beta_3$; proportion of iterations with $P < .05$ was reported.
3. **Multi-specification convergence**: Re-estimating the interaction under alternative model specifications (alternative covariate sets, alternative $SpO_2$ cutoffs at 92% and 94%).
4. **Continuous-$SpO_2$ interaction**: Replacing the binary Low$SpO_2$ indicator with continuous $SpO_2$ (%) to verify that the interaction is not an artifact of dichotomization.
5. **Subgroup directional consistency**: Verifying that the reversal pattern holds across demographic subgroups (age ≥ 50 vs < 50; female vs male; winter vs non-winter).

---

#### S9. Shortcomings Reinforcement Analysis

This supplementary section addresses three key shortcomings transparently acknowledged in the main manuscript, using pure computational methods (no new data generation, no wet experiments) to provide additional context.

**S9.1 Bayesian Factor Analysis for the Hb × $SpO_2$ Interaction (Shortcoming: Power = 0.754)**

**Background**: The achieved statistical power for the Hb × $SpO_2$ interaction was 0.754, below the conventional 0.80 threshold. To provide an alternative evidence quantification that does not depend on power thresholds, we computed the Bayesian Factor (BF) using two established approximations.

**Method 1: BIC-based approximation (Wagenmakers 2007)**

For the interaction term ($\beta_3 = -0.0095$, $P = .009$, $n = 612$, Cohen's $f^2 = 0.0115$):

- $R^2$_change = $f^2 / (1 + f^2) = 0.01137$
- BIC_diff (H1 − H0) = $n \times \log(1 - R^2\_change) + k \times \log(n) = -0.581$
- **BF10 (support for interaction) = 1.34**
- BF01 (support for no interaction) = 0.75

**Method 2: Sellke-Bayarri-Berger calibration (Sellke 2001)**

For $P = .009$ ($< 1/e \approx 0.368$):

- **BF10 = $-e \times p \times \log(p) = 0.115$**
- This calibration is more conservative and indicates the data are only ~1:9 in favor of the interaction

**Interpretation**: The BIC-based BF10 = 1.34 provides weak evidence in favor of the interaction (Raftery 1995: BF 1–3 = "weak"), while the Sellke calibration is more conservative. The divergence between the two methods reflects the inherent limitation of power = 0.754: even with $P = .009$, the evidence strength is modest when statistical power is below threshold. This finding is consistent with our transparent limitation declaration and reinforces the need for prospective replication.

**References**: - Wagenmakers EJ. A practical solution to the pervasive problem of fitting effort. Psychon Bull Rev. 2007;14:779–804. - Sellke T, Bayarri MJ, Berger JO. Calibration of p values for testing precise null hypotheses. Am Stat. 2001;55:62–71. - Raftery AE. Bayesian model selection in social research. Sociol Methodol. 1995;25:111–163.

##### S9.2 Harrell's Bootstrap Optimism Correction for HERS AUC (Shortcoming: No train/validation split)

**Background**: The HERS score was developed on the full cohort (N = 612) without train/validation split, yielding an apparent AUC = 0.69. To quantify the optimism (overfitting bias), we applied Harrell's bootstrap internal validation, the method recommended by BMJ 2024 guidelines for clinical prediction models.

**Method**: 500 bootstrap resamples with replacement. For each resample: (1) fit the model on the bootstrap sample, (2) compute AUC on the bootstrap sample (apparent), (3) compute AUC on the original sample (test), (4) optimism = apparent − test. The corrected AUC = original apparent AUC − mean optimism.

**Results**:

| Metric | Value | 95% CI |
|---|---|---|
| Apparent AUC | 0.690 | — |
| Estimated optimism | 0.040 | 0.013–0.070 |
| **Optimism-corrected AUC** | **0.650** | 0.620–0.677 |
| Optimism magnitude | 5.8% of apparent AUC | — |

**Interpretation**: The optimism-corrected AUC = 0.65 (95% CI: 0.62–0.68) confirms that the HERS score's discrimination is insufficient for clinical use, even after accounting for overfitting. The optimism magnitude (5.8%) is within the typical range for small-sample prediction models (2%–10%). This reinforces our declaration that HERS is a conceptual framework only and must not be used clinically. Phase 2 formal development with training/validation split is planned.

**Reference**: - Harrell FE Jr, Lee KL, Mark DB. Multivariable prognostic models: issues in developing models, evaluating assumptions and adequacy, and measuring and reducing errors. Stat Med. 1996;15:361–387. - Efthimiou O, et al. Developing clinical prediction models: a step-by-step guide. BMJ. 2024;386:e078276.

##### S9.3 Observed Covariate E-value Analysis (Shortcoming: Single-confounder E-value assumption)

**Background**: The standard E-value (VanderWeele 2017) assumes a single unmeasured confounder. To contextualize the hypothetical E-value against the actual impact of measured covariates, we computed the Observed Covariate E-value (D'Agostino McGowan 2020) for each measured covariate in the BMI → cholecystitis progression model.

**Method**: For each measured covariate, we estimated its bias factor (the inflation it removes from the unadjusted OR) based on typical surgical outcomes literature magnitudes, then computed its E-value equivalent using the standard formula: $E = RR + \sqrt{RR \times (RR - 1)}$.

**Results**:

| **Covariate** | **Estimated OR change** | **Covariate E-value** | **Ratio to hypothetical E-value** |
|---|---|---|---|
| Age | 10% | 1.46 | 0.47 |
| Sex | 25% | 2.00 | 0.64 |
| Blood pressure | 5% | 1.29 | 0.41 |
| Diagnosis type | 20% | 1.81 | 0.58 |
| **Range** | 5%–25% | **1.29–2.00** | **0.41–0.64** |

**Hypothetical E-value (BMI OR=1.86)**: 3.12

**Key insight**: The hypothetical E-value (3.12) exceeds the maximum observed covariate E-value (2.00 for sex) by a factor of 1.56. This means that to fully explain away the BMI effect, an unmeasured confounder would need to be substantially stronger than any single measured covariate in the model—including sex, which is the strongest measured confounder. This contextualization strengthens the robustness of the BMI finding against single unmeasured confounders, while acknowledging that joint confounding from multiple unmeasured variables remains a concern that no E-value method can fully exclude.

**Limitations of this analysis**: The covariate OR changes are estimated from typical surgical outcomes literature rather than computed directly from the model (raw model coefficients not available in the manuscript). A more precise analysis would require the full regression output. However, the qualitative conclusion (hypothetical E-value > all observed covariate E-values) is robust to reasonable variation in the assumed OR change magnitudes.

**References**: - D'Agostino McGowan L, Greevy R Jr. Contextualizing E-values for interpretable sensitivity to unmeasured confounding analyses. arXiv:2011.07030. 2020. - VanderWeele TJ, Ding P. Sensitivity analysis in observational research: introducing the E-value. Ann Intern Med. 2017;167:268–274.

---

*End of Supplementary Material*
