## Supplementary material for "Patient-Specific Adaptations in ERAS for High-Altitude Laparoscopic Cholecystectomy: The PAERS Hypothesis": STROBE Checklist

### STROBE Checklist — Paper 5

**Manuscript**: From "One-Size-Fits-All" to Precision: Patient-Specific Adaptations in ERAS Redefine Perioperative Risk Stratification for High-Altitude Laparoscopic Cholecystectomy — The PAERS Hypothesis **Reporting Guideline**: STROBE (Strengthening the Reporting of Observational Studies in Epidemiology) — Observational Cohort Study with Hypothesis-Generating Integration **Checklist Version**: STROBE 2007 (updated 2014) **Last Updated**: 2026-08-21

#### Checklist

| Item | Recommendation | Page/Section | Reported |
|---|---|---|---|
| **Title and abstract** | | | |
| 1 | (a) Indicate study design with a commonly used term in the title or abstract | Abstract (Methods) | ✅ |
| 1 | (b) Provide in the abstract an informative and balanced summary of what was done and what was found | Abstract | ✅ |
| **Introduction** | | | |
| 2 | Explain the scientific background and rationale for the investigation being reported | Introduction | ✅ |
| 3 | State specific objectives, including any prespecified hypotheses | Introduction (last paragraph); Methods 2.1 (post hoc hypothesis formulation transparently declared) | ✅ |
| **Methods** | | | |
| 4 | Present key elements of study design early in the paper | Methods (Study Design) | ✅ |
| 5 | Describe the setting, locations, and relevant dates, including periods of recruitment, exposure, follow-up, and data collection | Methods (Setting: Qinghai Red Cross Hospital, 2260 m, 2018-2023) | ✅ |
| 6 | (a) Give the eligibility criteria, and the sources and methods of selection of participants | Methods (Participants: 612 adults, elective LC) | ✅ |
| 7 | Clearly define all outcomes, exposures, predictors, potential confounders, and effect modifiers | Methods (Variables: BMI, Hb, $SpO_2$, surgeon, EPAS1 carrier inference) | ✅ |
| 8 | For each variable of interest, give sources of data and details of methods of assessment | Methods (Data Sources: HIS, laboratory, surgical records) | ✅ |
| 9 | Describe any efforts to address potential sources of bias | Methods (Bias: multivariable adjustment, E-value sensitivity analysis, 5 robustness analyses) | ✅ |
| 10 | Explain how the study size was arrived at | Methods (Sample Size: 612 patients, post-hoc power 0.754 transparently reported) | ✅ |
| 11 | Explain how quantitative variables were handled in the analyses. If relevant, describe which groupings were chosen and why | Methods (Statistical Analysis: BMI strata, $SpO_2$ strata, Hb threshold 180 g/L) | ✅ |
| 12 | (a) Describe all statistical methods, including those used to control for confounding | Methods (Statistical Analysis: multivariable logistic/linear regression, RCS, interaction | ✅ |

| Item | Recommendation | Page/Section | Reported |
|---|---|---|---|
| | | models) | |
| 12 | (b) Describe any methods used to examine subgroups and interactions | Methods (Tier 2: Hb × $SpO_2$ interaction; Tier 3: genetic modifier inference) | ✅ |
| 12 | (c) Explain how addressing missing data was addressed | Methods (Missing data: excluded cases with incomplete records, n=38 from initial 650) | ✅ |
| 12 | (d) If applicable, describe analytical methods for cohort study (e.g., loss-to-follow-up) | Methods (No loss-to-follow-up; complete 30-day follow-up) | ✅ |
| **Results** | | | |
| 13 | (a) Report numbers of individuals at each stage of the study—e.g., potentially eligible, examined for eligibility, confirmed eligible, included in the study, completing follow-up, and analysed | Results (Study population: 650 screened, 612 analyzed; Figure 1) | ✅ |
| 13 | (b) Give reasons for non-participation at each stage | Results (Figure 1: 38 excluded for incomplete data) | ✅ |
| 13 | (c) Consider use of a flow diagram | Figure 1 (Study Flow Diagram) | ✅ |
| 14 | (a) Give characteristics of study participants and information on exposures and potential confounders | Table 1 (Baseline characteristics by BMI stratum) | ✅ |
| 14 | (b) Indicate number of participants with missing data for each variable of interest | Table 1 footnote (zero missing data after exclusion) | ✅ |
| 15 | Report numbers of outcome events or summary measures | Results (3.1-3.5: LOS, complications, OR, β, F, ρ) | ✅ |
| 16 | (a) Give unadjusted estimates and, if applicable, confounder-adjusted estimates and their precision (e.g., 95% confidence interval) | Results (Tables 2-3; unadjusted and adjusted OR/β with 95% CI) | ✅ |
| 16 | (b) Report category boundaries when continuous variables were categorized | Methods (BMI strata: 18.5-23.9 normal, 24-27.9 overweight, ≥28 obese; $SpO_2$: <93%, 93-95%, ≥96%) | ✅ |
| 16 | (c) If relevant, consider translating estimates of relative risk to absolute risk for a meaningful time period | Results (LOS in days as absolute measure) | ✅ |
| 17 | Report other analyses done—e.g., analyses of subgroups and interactions, and sensitivity analyses | Results (3.3: Hb × $SpO_2$ interaction; 3.4: 5 robustness analyses; Supplementary S7: E-value) | ✅ |
| **Discussion** | | | |
| 18 | Summarise key results with a focus on their relevance to the objectives | Discussion (4.1 Key Findings) | ✅ |
| 19 | Discuss possible limitations of the study, including sources of potential bias or imprecision, and the adequacy of the study to address the objectives | Discussion (4.5 Limitations L1-L7: single-center, single altitude, genetic inference, statistical power, HERS conceptual, no rheology data, LOS composite) | ✅ |
| 20 | Give a cautious overall interpretation of results considering objectives, limitations, and multiplicity of analyses | Discussion (4.4 PAERS hypothesis as conditional extension, not negation; honest declaration of hypothesis-generating positioning) | ✅ |
| 21 | Discuss the generalizability (external validity) of the study results | Discussion (4.6 Future Directions: Phase 2/3 multi-center validation planned) | ✅ |

| Item | Recommendation | Page/Section | Reported |
| --- | --- | --- | --- |
| **Other information** | | | |
| 22 | Give the source of funding and the role of the funders | Declarations (Funding: YNZXKT2026009, acquired by Zhiqiang Wang; funder had no role) | ✅ |
| 23 | Give approval number and the ethical committee judgment | Declarations (Ethics: LW-2026-73, IRB Qinghai Red Cross Hospital, waiver of informed consent) | ✅ |

#### Additional Reporting Items (Hypothesis-Generating Study)

| Item | Recommendation | Page/Section | Reported |
| --- | --- | --- | --- |
| H1 | Transparently declare post hoc hypothesis formulation | Methods 2.1 (PAERS hypothesis formulated post hoc through integration of four interconnected analyses) | ✅ |
| H2 | Provide falsifiable predictions for prospective validation | Discussion 4.6 (F1-F3 predictions with validation methods and falsification consequences) | ✅ |
| H3 | Distinguish empirical findings from inferential framework | Throughout (Dimension 3 explicitly labeled as "population-level inference, not individual genotyping"; HERS labeled as "conceptual framework only") | ✅ |
| H4 | Report E-values for unmeasured confounding | Supplementary S7 (E-values for all 6 main findings) | ✅ |
| H5 | Disclose Phase 1 positioning | Methods 2.1 (hypothesis-generating, single-center); Discussion 4.6 (Phase 1/2/3 roadmap) | ✅ |

#### Transparency Declaration

This study adheres to FAIR compliance principles, transparently disclosing: 1. **Negative results**: ICGC cohort negative findings for EPAS1→STC2 correlation (ρ=−0.051) reported transparently 2. **Limitations**: 7 explicit limitations (L1-L7) with severity grading 3. **Statistical power**: Achieved power 0.754 (<0.80) transparently reported and discussed 4. **Genetic inference nature**: All genetic modification analyses explicitly labeled as "population-level inference, not individual-level empirical findings" 5. **HERS score status**: Conceptual framework only, not externally validated, must not be used clinically 6. **Preprint status**: This manuscript is a preprint and has not been peer-reviewed

*End of STROBE Checklist for Paper 5*
